# AUTISM SPECTRUM DISORDER: A POSSIBLE LINK WITH CHONDROITIN SULFATE

**DOI:** 10.1101/2020.10.15.20212910

**Authors:** Eduardo Listik, Marcia Listik, Clarice Listik, Leny Toma

**Author notes:** Corresponding author: Eduardo Listik, Ph.D., WTI 330F, 1824 Sixth Avenue South, Birmingham, AL, 35233., /1. Authors’ contributions: ED Prepared all samples, performed data analysis, and drafted the initial manuscript. ML Collected data, carried out the patient care and selection. LT Performed gel electrophoresis. All authors reviewed and revised the manuscript.

## Abstract

This study aimed to determine discrepancies in the urinary glycosaminoglycan profiles of autism spectrum disorder (ASD) patients (n=9) when compared with those from healthy volunteers (HVs, n=3). The guardians and/or educators for each participant also returned a validated Autism Behavior Checklist (ABC). The urinary chondroitin sulfate (CS) concentration was 46.1% lower in the ASD group than in the HV group. The ABC score and the urinary CS concentration were negatively correlated (Spearman’s ρ=– 0.2635), indicating that as the severity of the clinical aspect of this disorder increased, the urinary CS concentration decreased. These results suggest that low CS concentrations in the urine may be associated with ASD, and could be measured using a fast and low-cost method for diagnostics.

## INTRODUCTION

Autism spectrum disorder (ASD) are characterized by a lack of social interaction and impaired verbal and non-verbal communication and can be diagnosed as early as three years of age (Park et al., 2016).

Over the last two decades, ASD diagnoses have steadily increased (Fisch, 2012). Some specialists have argued that this increase has been due to an expansion of the diagnostic criteria and increased social awareness; however, newly diagnosed patients appear to have similar profiles as those diagnosed using previous assessments, and the new criteria do not seem to have influenced these results (Baio et al., 2018; Graf, Miller, Epstein, & Rapin, 2017). The diagnosis of ASD is observational and based on the patient’s behavior, although some biomarkers have been proposed (Galiana-Simal, Munoz-Martinez, Calero-Bueno, Vela-Romero, & Beato-Fernandez, 2018).

Glycosaminoglycans (GAGs) are complex polysaccharides, composed of repeating disaccharide units. The composition of these units and their linking patterns result in the formation of distinct GAGs, such as chondroitin sulfate (CS), dermatan sulfate (DS) and heparan sulfate (HS) (Gandhi & Mancera, 2008). GAGs may conjugate with proteins, known as proteoglycans, and are essential for embryogenesis and neuronal migration during development. CS proteoglycan mutations may be associated with schizophrenia and bipolar disorder (Couchman, 2010; Iozzo & Schaefer, 2015; Maeda, 2015; Muhleisen et al., 2012; Nakato & Li, 2016; Turner & Grose, 2010). Additionally, excreted GAGs in the urine may be used to diagnose mucopolysaccharidoses (MPS) (Coutinho, Lacerda, & Alves, 2012). Recently, the genes associated with glycan synthesis have been thought to play a role in the development of ASD (Dwyer & Esko, 2016; Endreffy, Bjorklund, Dicso, Urbina, & Endreffy, 2016). Therefore, this study aimed to assess whether the concentrations of GAGs in the urine of ASD patients can be used to facilitate the diagnosis of ASD, potentially resulting in a better understanding of the ASD pathophysiology and possible therapeutic targets.

## METHODS

### Standard protocol approvals, registrations, and patient consents

All procedures performed in studies involving human participants were in accordance with the ethical standards of the Institutional Ethics Committee of Research (n° 14667219.0.0000.5505) and with the 1964 Helsinki declaration and its later amendments or comparable ethical standards. Written informed consent/assent was obtained from all participants or their guardians.

### Patients and clinical assessments

A pediatric neurologist assessed patients with behavioral or communication difficulties for the evaluation of a possible ASD diagnosis. Exclusion criteria were as follows: younger than 2 years; older than 8 years; neurological lesions; mental deficiency; and other diseases. Patients were also excluded if additional cases of ASD or consanguinity were detected in their family histories. The same exclusion criteria were applied to the healthy volunteers (HVs). The guardians and/or educators for all participants were asked to respond to an electronic version of the validated Autism Behavior Checklist (ABC), which is often used for its ability to identify children with autism. (Marteleto & Pedromonico, 2005) Both total scores and sub-scores were calculated. The sub-scores included: sensory stimulus (SS), use of body and objects (BO), language (LG), and personal-social development (PS). All participants had a urine sample collected, which was stored at −20 °C until processing.

### Glycosaminoglycan analysis

Urine samples were centrifuged at 5,000 rpm for 10 min, 4 mL of the supernatant were concentrated, using Amicon^®^ Ultra centrifugal filters with a molecular weight cut-off of 3 kDa (Millipore, Massachusetts, USA), and the concentrate was desalted using 10 mM NaCl (Sun et al., 2015).

Concentrated samples were applied to 0.6% agarose slabs, in diaminopropane-acetate, 0.05 M (pH 9.0) buffer, and the GAGs were electrophoretically separated at 100 V for 60 min (Dietrich & Dietrich, 1976; Jaques, Ballieux, Dietrich, & Kavanagh, 1968). Gels were maintained in cetyltrimethylammonium bromide at 0.2% for 1 h and dehydrated completely before being stained with a solution of 0.1% toluidine blue, 1% acetic acid and 50% ethanol for 20 min. The excess dye was removed with quick washes of a destaining solution containing 1% acetic acid and 50% ethanol. The GAG bands were densitometrically quantified and compared with an HS/DS/CS standard to calculate the GAG concentrations in the urine.

### Data analysis

GAG concentrations and ABC scores were compared between the HV and ASD groups using a Mann-Whitney test, with p < 0.05 representing a significant difference. The two variables were also examined for a possible correlation using Spearman’s rank-order correlation test. Variables were screened for outliers using the robust regression and outlier removal test (ROUT’s test).

### Data availability Statement

All data associated with this study are available from the corresponding author, upon reasonable request.

## RESULTS

This study included 12 participants, nine who were diagnosed with ASD and three HVs. One ASD patient was excluded based on the ROUT’s test results for the ABC scores, and one HV was excluded due to sample deterioration. The sample descriptors are summarized in Table 1.

**Table 1.** Sample descriptors for ASD patients and HVs. The sample size, mean age, distribution of sex, and ABC sub-scores and total score are shown for each group. The number of individuals removed from each group is shown in the parentheses.

| Descriptor | ASD patients | HV |
| --- | --- | --- |
| Sample size | 9 (-1) | 3 (-1) |
| Mean age | 3.9 ± 1.1 years | 8.0 ± 0.0 years |
| Sex | All male | All male |
| Ethnicity | Caucasian (88.9%)<br>Asian (11.1%) | Caucasian (100%) |
| ABC sensory stimulus (SS) | 8.6 ± 4.4 | 0.0 ± 0.0 |
| ABC relationship (RE) | 10.6 ± 7.2 | 1.5 ± 2.1 |
| ABC use of body and objects (BO) | 14.5 ± 8.4 | 0.0 ± 0.0 |
| ABC language (LG) | 16.2 ± 6.0 | 0.0 ± 0.0 |
| ABC personal and social development<br>(PS) | 13.7 ± 6.0 | 3.5 ± 0.7 |
| ABC total score | 63.5 ± 23.4 | 5.0 ± 2.8 |

Our data revealed that the ABC scores for the ASD patients (63.5 ± 23.4) were significantly higher than those for the HVs (5.0 ± 2.8), as shown in Figure 1B. The ABC sub-scores revealed that ASD patients received higher scores for all investigated parameters, except for the relationship (RE) category (Figure 1B). The gel electrophoresis for the assessment of GAG concentrations in the urine showed the prevalence of CS, small signs of HS and none DS (Figure 1A) A significant reduction (p = 0.04) of 46.1% in the CS concentrations was observed in the urine of ASD patients compared with the urine of HV patients. Whereas ASD patients had a mean CS concentration of 9.44 ± 5.75 mg/L, HVs had a considerably higher mean concentration of 17.51 ± 4.04 mg/L (Figure 1C).

**Figure 1.**
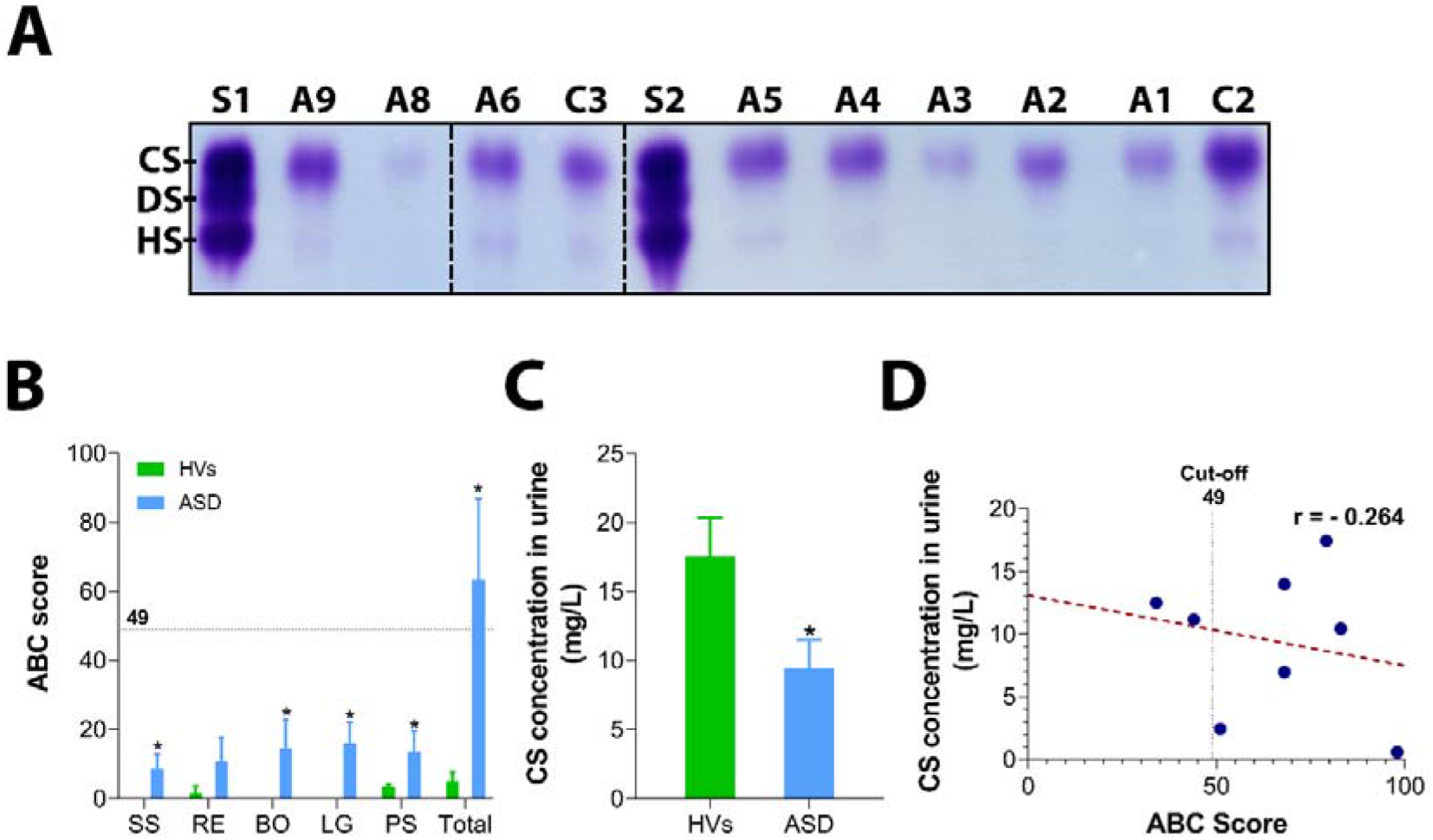
Discrepancies in the CS concentrations in the urine between ASD patients and HVs. (A) Processed urine samples were analyzed for GAG concentrations using an electrophoretic method, in which CS was prevalent. S1 and S2 are the CS/DS/HS standards, A1 – A9 (except A7 which was excluded) are ASD patients, and C2 and C3 are HVs. (B) ASD patients displayed significantly higher ABC scores than HVs. All of the sub-scores, except for relationship (RE), were higher in ASD patients than in HVs. The horizontal dotted line at *y* = 49 indicates the scale cut-off. Other sub-scores abbreviations: sensory stimulus (SS), use of body and objects (BO), language (LG), and personal-social development (PS). (C) The measured CS concentration in the urine of ASD patients is significantly lower than that in HVs. (D) A negative correlation between ABC scores and CS concentration in urine, based on the Spearman’s ρ.

Although a larger sample size would be necessary to estimate a precise correlation between the two examined variables (i.e., ABC score *vs*. CS concentration in urine), we observe that ABC scores for ASD patients were negatively correlated (Spearman’s ρ = –0.2635) with the measured GAG concentrations in the urine, which could indicate that lower levels of CS in the urine may be associated with worse ASD clinical diagnoses.

## DISCUSSION

As previously mentioned, GAGs have been used to monitor MPS. During these pathologies, CS, DS, HS or keratan sulfate (KS) can be observed in patient urine samples, depending on the MPS (Coutinho et al., 2012). However, we discovered that ASD patients display lower urinary CS concentrations for reasons that remain unknown.

There is no corroborating evidence in the literature that would explain the lower CS concentrations observed in the urine samples from ASD patients. The deletion of *EXT1, GPC5, GPC6*, and mutation of *HS3ST5* and *SGSH*, appear to be associated with ASD (Dwyer & Esko, 2016). These genes participate in HS synthesis, sulfation, degradation, or HS proteoglycan synthesis. In addition, the deletion of *B3GALT6*, a gene that affects both HS and CS synthesis, also appears to be implicated with ASD (van der Zwaag et al., 2009).

The detection of GAGs in the urine may not be appropriate for the detection of HS in these samples, as this GAGs was hardly detectable either in patients with ASD or HVs urines; however, because the glycan balance may be altered in patients with ASD, these alterations can be verified based on the CS concentrations, which revealed significant discrepancies in this pilot study. A larger sample may establish whether urinary CS concentrations can be utilized as a biomarker for ASD and help shed light on the still unknown ASD pathophysiology, and this is a clear limitation of this study. In addition, more studies with HS or specific proteoglycans in the blood or other samples may also increase our understanding of ASD pathophysiology.

In this pilot study, we were able to infer that ASD patients may excrete less CS in the urine, which can be assessed using a fast and low-cost analytical method.

